# Wellbeing After Stroke-2 (WAterS-2): a feasibility study with process evaluation exploring inclusive, accessible, online psychological support after stroke

**DOI:** 10.64898/2026.06.12.26355528

**Authors:** Verity Longley, Kate Woodward-Nutt, Sarah Cotterill, Niki Chouliara, Shirley Thomas, Ann Bamford, Audrey Bowen, Emma Patchwood

## Abstract

**Objectives:** Explore feasibility and acceptability of upskilling a workforce to deliver a co-developed intervention, based on Acceptance and Commitment Therapy (ACT), to support psychological adjustment post-stroke targeting underserved groups.

**Design:** Multi-site, single-arm feasibility study with embedded mixed-methods process evaluation (ISRCTN17628580).

**Setting:** Four NHS community stroke services across England.

**Participants 1:** Stroke survivors ≥18 years of age, ≥4 months post-stroke, reporting psychological difficulties adjusting to stroke, able to consent and access remote group sessions in English; 2. Group facilitators from NHS stroke services, not ACT specialists.

**Intervention:** WAterS-2: an eight-session, remotely-delivered ACT-informed group intervention.

**Outcome measures:** Recruitment, fidelity, safety, acceptability and perceived value were assessed using fidelity checklists, post-intervention surveys and semi-structured interviews with stroke survivors and facilitators. Clinical outcomes including mood (HADS), wellbeing (ONS4), psychological flexibility (AAQ-ABI), measured post-group and three-months later.

**Results:** Nineteen stroke survivors recruited (mean 9.6 months post-stroke; n=5 (26%) minoritised ethnicities; n=10 (52%) with aphasia). Thirteen facilitators - including two peer support workers - delivered the intervention with fidelity following structured training across four services. Drop-out was low (2/19; 11%); with 15 (79%) attending ≥5/8 sessions. Remote data collection was feasible (79% follow-up completion), with no adverse events recorded. Acceptability was high: survivors valued peer connection, grounding and mindfulness practices. ACT metaphors were helpful for some but challenging for others, including some with aphasia. Online delivery was suitable but limited informal connection. Facilitators reported increased capability, incorporating ACT skills into routine care. NHS workforce pressures and geographically-constrained referral pathways limited recruitment reach.

**Conclusions:** WAterS-2 is feasible, safe, acceptable and inclusive. A mixed workforce, including NHS peer support workers, can be upskilled to deliver with fidelity. Inclusion of underserved groups is achievable but requires active strategies beyond standard NHS referral routes. Findings inform a provisional logic model and a future pragmatic trial.

**Registration:** ISRCTN17628580 (registered 16/08/2024).

**Strengths and Limitations of this study:**

- This feasibility study used a mixed-methods process evaluation, enabling a rich account of both feasibility indicators and implementation considerations across NHS sites.
- We included stroke survivors with communication difficulties and from minoritised ethnic backgrounds, groups typically excluded from stroke rehabilitation research.
- People and Communities involvement was embedded throughout all stages of study design, delivery and interpretation.
- The single-arm design and small sample size prevent causal or effectiveness inferences, and delivery of only one group per site limits assessment of variability across teams.
- Recruitment was undertaken by clinical staff from existing caseloads, introducing potential for referral bias.

## Introduction

Supporting psychological adjustment is a core component of stroke rehabilitation and has remained a national research priority in the UK since 2012 [1, 2]. However, stroke survivors often report feeling their psychological needs are unmet [3].

These unmet needs reflect wider health inequalities [4] across the stroke pathway. For example, people in the UK from minoritised ethnic communities have a higher risk of stroke and poorer outcomes than White British populations [5]. Communication difficulties such as aphasia are common after stroke and are associated with worse recovery [6]. These inequalities extend to psychological wellbeing: both groups are more likely to experience ongoing mental health difficulties after stroke and face barriers to accessing appropriate support [7-10]. Psychological support within stroke services needs to be evidence-based, inclusive and accessible.

Acceptance and Commitment Therapy (ACT) is a third-wave, transdiagnostic cognitive-behavioural psychotherapy, offering a promising model for supporting psychological adjustment after stroke [11]. ACT aims to improve psychological flexibility — supporting individuals to engage in meaningful activities despite ongoing difficulties — rather than focusing solely on symptom reduction. There is growing evidence supporting ACT’s efficacy in improving psychological wellbeing and preventing depression post-stroke [12, 13]. Additionally, the latest National Clinical Guideline for Stroke for the UK and Ireland recommended ACT for stroke survivors with anxiety or depression [14].

Many stroke services in the UK have limited access to psychology staff. [15]. One potential solution is task-sharing, where non-specialist staff are trained to deliver structured psychological interventions with appropriate supervision [16]. This approach is supported by global mental health evidence and World Health Organization guidance [17, 18], but has had little testing within UK stroke rehabilitation services. Remotely delivered interventions (telerehabilitation), may help optimise service delivery by improving reach and access in a resource efficient way [19, 20], especially in a post-pandemic world [21].

The Wellbeing After Stroke (WAterS) study was developed to address these gaps. In WAterS-1 we co-developed an online, group-based ACT-informed intervention, feasibility tested in the stroke-specialist charitable third sector [22]. It showed promising feasibility and acceptability but did not test delivery within NHS services nor address inequalities for underserved groups.

WAterS-2 refers to the subsequently enhanced intervention; refined through more co-development, including more diverse public involvement and interviews with professional stakeholders [23]. Refinements included fewer sessions and adaptation of the content to be more accessible to underserved groups. We aimed to evaluate the feasibility of delivering WAterS-2 online, in groups within NHS stroke services, through workforce upskilling, and recruitment of stroke survivors with communication difficulties and those from minoritised ethnic communities. Our five objectives were to:

1. assess whether a non-psychology NHS stroke workforce could be upskilled (trained and supported to deliver WAterS-2 online groups with fidelity and safety);
2. examine the feasibility of identifying, recruiting and retaining marginalised stroke survivors, focused on people with aphasia and those from minoritised ethnic backgrounds;
3. evaluate the feasibility of collecting outcome data remotely at multiple timepoints;
4. explore the acceptability and perceived value of WAterS-2 for stroke survivors and staff facilitators;
5. identify contextual and implementation factors likely to influence future delivery in routine NHS practice

## Methods

This was a multi-site, single-arm feasibility study with embedded quantitative and qualitative process evaluation. The study was informed by the Medical Research Council framework for developing and evaluating complex interventions [24] and by ongoing collaborative involvement from People and Communities contributors [25, 26].

Ethics approval was granted by Wales Research Ethics Committee-6 (REC ref: 24/WA/0238; IRAS ID: 287785). The study and protocol were prospectively registered [27]. Study quality and conduct was monitored by a Management Group and an external Steering Committee. The study ran from January 2025 to December 2025 with recruitment taking place between February and May 2025.

### Involving People and Communities

We embedded People and Communities Involvement using what we call a ‘Core and Cluster’ approach. Our full involvement strategy is available [28]. The core Research Advisory Panel (RAP) included seven individuals with a range of experiences including stroke, communication difficulties, community advocacy, and minoritised ethnicity. The RAP was led by a lay chair (co-author ABa) and met frequently, contributing to all aspects of the study from design and materials through to interpretation and dissemination. Wider, flexible engagement with community organisations and connectors (clusters’) extended broader reach of involvement. Contributors were reimbursed for their time and expenses.

### Setting and Participants

NHS community stroke services across England were eligible to participate. We recruited and trained staff as group facilitators. Staff were eligible if they worked within participating services and could attend training; prior experience with ACT was not required.

Stroke survivors were eligible to participate in groups if they were over 18 years old, at least four months post-stroke (no upper limit), reported ongoing difficulties psychologically adjusting to life after stroke, and were able to give informed consent to participate in a remotely delivered group in English. Exclusion criteria included high clinical risk requiring specialist psychological support or inability to provide informed consent. We used accessible recruitment materials and supportive communication strategies to recruit individuals with communication difficulties.

Informed consent was obtained from all participants by the research team. Stroke survivors could request a supporting individual (e.g., family members or friends) to assist with engagement.

Group facilitators were opportunistically identified through participating service leads. Stroke survivors were primarily identified and referred by clinical teams using existing caseloads; teams were asked to target stroke survivors from minoritised ethnic background and/or with communication difficulties. Planned supplementary recruitment routes included community advertisements and self-referral, as per the original WAterS [22].

### Facilitator Training

Facilitators completed four half-day online training sessions delivered by clinical neuropsychologists with expertise in ACT and stroke care. Training included introduction to ACT principles, modelling of group processes, rehearsal of session content, and practice using the manual. Weekly online group supervision was available during delivery.

### Intervention: group support

Groups began once sufficient stroke survivor participants were enrolled locally at each site. Appendix 1 provides detailed description of the intervention to support replication [29]. WAterS-2 is an eight-week, eight-session remotely-delivered group intervention for 4-10 participants informed by ACT and neurorehabilitation principles. Sessions last 120 minutes delivered via Microsoft Teams, co-delivered by two trained facilitators, with rest breaks.

Sessions combined psychoeducation, experiential ACT-based exercises (e.g., mindfulness, acceptance, values clarification and committed action), and structured home practice, supported by a participant handbook and visual materials. A highly-structured manual and scripts were used to support consistency and confidence among facilitators.

### Data Collection

We collected all data remotely using telephone, video conferencing (Microsoft Teams) and anonymous online surveys (Qualtrics). Baseline demographic and clinical information was collected from participants following consent. We collected data at baseline, immediately post-group end, and three-months later (stroke survivors only). See Table 1 for a summary of measures and timepoints.

**Table 1.** – Summary of remote data collection by participant group and timepoint

| Participant group | Measure / data source | Baseline | Post training | During groups | Post 8 week group end (Time 1) | 3-months after Time 1 |
| --- | --- | --- | --- | --- | --- | --- |
| **Stroke survivors** | Demographic and clinical characteristics (stroke severity SmRS) | X |  |  |  |  |
|  | Communication and cognitive screening (FAST, QAB, TOMS) | X |  |  |  |  |
|  | Mood (HADS) | X |  |  | X | X |
|  | Wellbeing (ONS4) | X |  |  | X | X |
|  | Health-related quality of life (EQ-5D-5L) | X |  |  | X | X |
|  | Psychological flexibility (AAQ-ABI) |  |  |  | X | X |
|  | Values-based living (VQ) |  |  |  | X | X |
|  | Brief feedback survey |  |  |  | X |  |
|  | Semi-structured interview |  |  |  | X |  |
| **Facilitators** | Demographics and professional background | X |  |  |  |  |
|  | Training feedback survey |  | X |  |  |  |
|  | Delivery feedback survey |  |  |  | X |  |
|  | Fidelity checklist (session components delivered, timing, attendance) |  |  | X (each session) |  |  |
|  | Semi-structured interview |  |  |  | X |  |

Remotely collected data included:

- Feasibility indicators: referral and consent rates, retention (e.g. attendance at ≥5 of 8 sessions), and outcome completeness.
- Stroke survivor baseline characteristics: demographic information and brief clinical indicators: Frenchay Aphasia Screening Test (FAST)[30]; Quick Aphasia Battery for Comprehension (QAB)[31]; simplified modified Rankin Scale (SmRS)[32]. Following Teams calls and clinical indicators, researchers rated stroke survivor participants on the therapy outcome measures (TOMS) [33] scales for aphasia (activity level) and cognition (impairment level).
- Facilitator baseline characteristics: demographic information; professional background / role; years of experience.
- PROMS for stroke survivors (see Table 1): Hospital Anxiety and Depression Scale (HADS) [34], the Office for National Statistics Personal Wellbeing questions (ONS4) [35] to assess subjective wellbeing, and the EQ-5D-5L [36] to assess health-related quality of life and utility values. ACT-consistent process PROMS included the Acceptance and Action Questionnaire – Acquired Brain Injury (AAQ-ABI) [37], assessing psychological flexibility, and the Valuing Questionnaire (VQ) [38], assessing engagement with personally meaningful activities.
- Fidelity indicators: attendance records, session duration and structured checklists completed by facilitators after each session, as used in previous research [39]. (See example fidelity checklist in Appendix 2).
- Acceptability and perceived value: explored using brief surveys and qualitative semi-structured interviews with facilitators and stroke survivors addressing experiences of delivery, usefulness, accessibility, perceived benefits and limitations (See Interview topic guides in Appendix 3).
- Safety: monitored via recording of adverse events by clinical team, and ongoing participant self-report.

### Analysis

We summarised quantitative data descriptively using SPSS. Categorical variables were reported as counts and percentages. Continuous variables were reported as mean (standard deviation) and/or median (interquartile range), appropriate to their distribution. For repeated measures, change scores with 95% confidence intervals were presented for participants with paired data. Measures were scored according to published guidance where available. Where guidance was absent, missing items were imputed with the within-person median if ≥80% of items were present; otherwise treated as missing. These procedures supported assessment of data completeness. No hypothesis testing for intervention effectiveness was powered or planned.

Qualitative interviews were audio-recorded, transcribed verbatim and analysed using a structured framework approach [40] aligned with study objectives. Coding was undertaken in NVivo independently by EP, KW-N and VL, then discussed, grouped and refined using an iterative process to enhance credibility and consistency. We considered findings from quantitative and qualitative sources together during interpretation and reporting to provide an account of feasibility, acceptability and implementation considerations.

## Results

Figure 1 summarises study processes. Four sites participated; 13 facilitators were recruited and completed training. Clinical teams recorded 63 stroke survivors on screening logs, of whom 31 (49%) consented to contact from the research team, and 19 of 31 (61%) consented to participate. Reasons recorded for non-referral or decline included limited technology access, language barriers (all content was delivered in English), preference not to engage in group-based psychological support, and perceived mismatch with individual needs (e.g., mental health or cognitive difficulties considered too severe for group participation). In contrast to WAterS-1, none of the planned supplementary routes to recruitment were utilised e.g. self-referral, due to NHS team concerns about duty of care for individuals not already on their caseload (current or historic).

**Figure 1.**
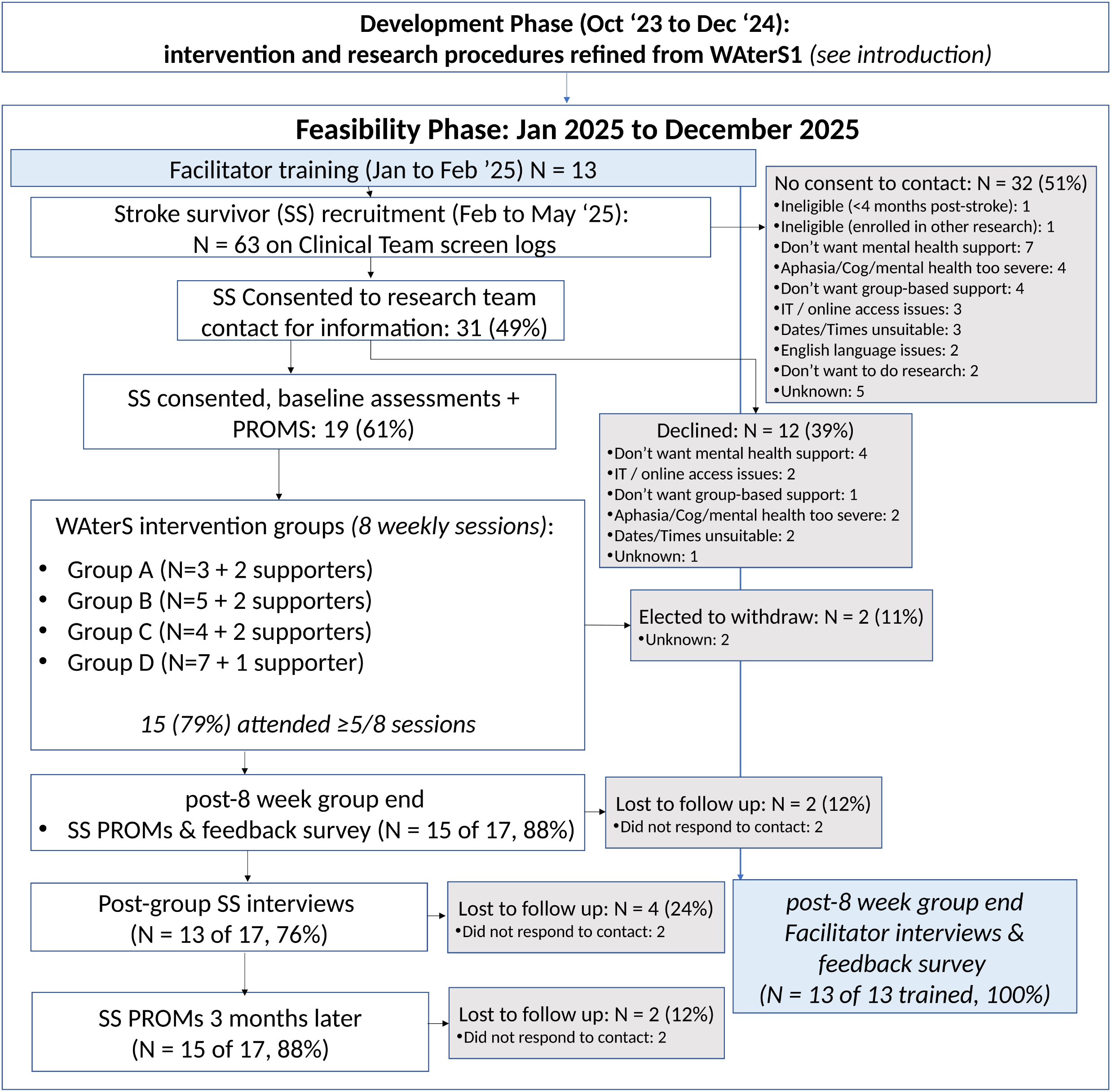
Study timelines and flow of participants.

Each site delivered one group during the study period. Nineteen stroke survivors attended across the four groups. Stroke survivor baseline characteristics are shown across Table 2 and Table 3. The mean age of participants was 57.5 years (SD 15.4; range 23–81), 62% were male, and the mean time since stroke was 9.6 months (SD 8.4). Baseline mood scores on the HADS suggest that 7 (37%) were experiencing anxiety and 9 (47%) were experiencing depression (defined as score of 11 or more on each subscale). Fourteen (74%) were classed as “dependent” on SmRS (score of 3 or more). Six supporting individuals (family members) were requested by stroke survivors and were invited to sessions to support engagement. Two stroke survivors (11%) withdrew during group delivery, attending 2 and 3 sessions; 15 (79%) completed the therapy, defined as attending at least five of eight sessions; and 2 (11%) attended four sessions or fewer. Outcome data were returned by 15 (79%) participants post-group and at three-month follow-up, with 13 (68%) stroke survivors agreeing to interview.

**Table 2.**
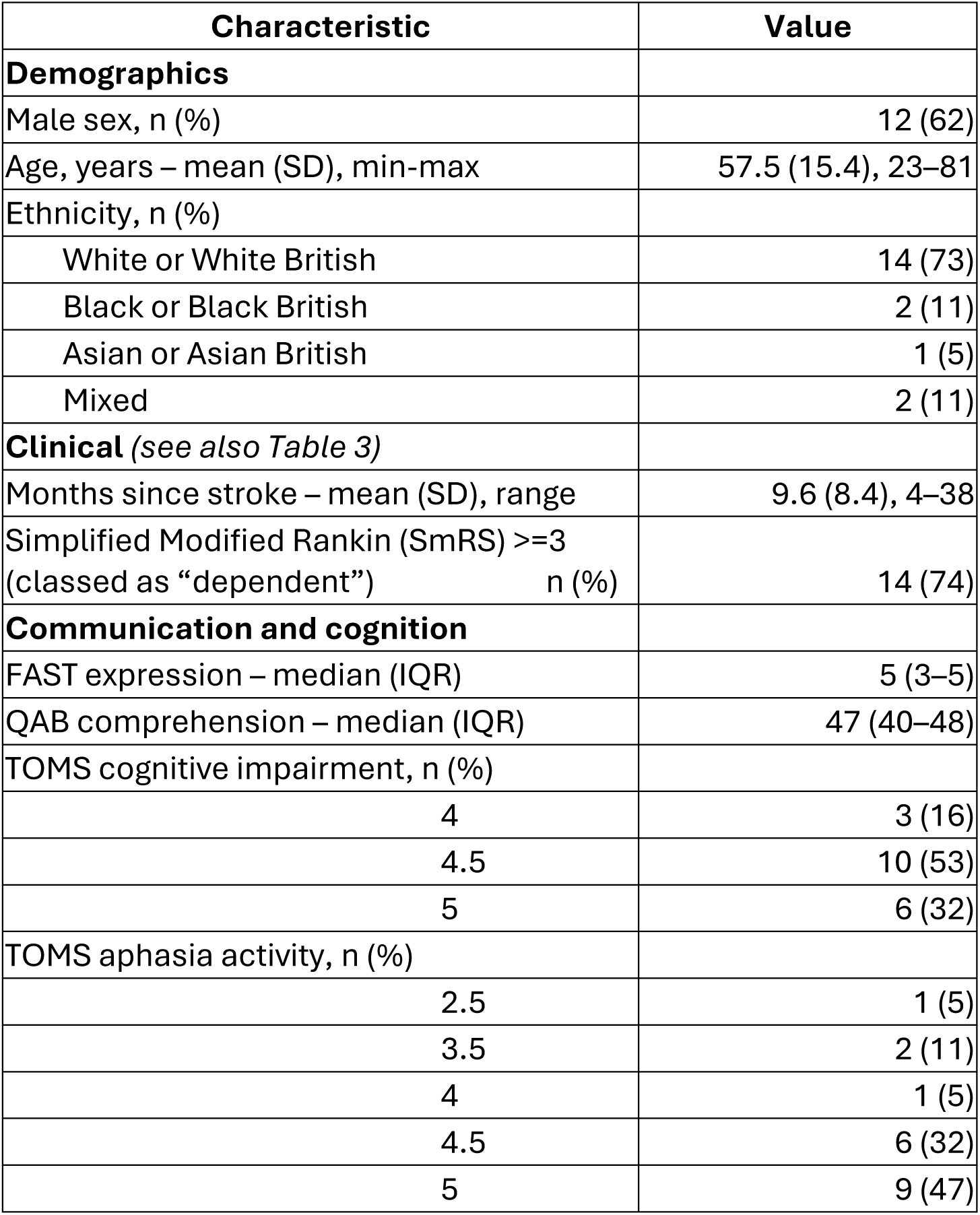
Baseline profile of consented stroke survivor participants (N=19)

| Characteristic | Value |
| --- | --- |
| **Demographics** |  |
| Male sex, n (%) | 12 (62) |
| Age, years – mean (SD), min-max | 57.5 (15.4), 23–81 |
| Ethnicity, n (%) |  |
| White or White British | 14 (73) |
| Black or Black British | 2 (11) |
| Asian or Asian British | 1 (5) |
| Mixed | 2 (11) |
| **Clinical** (see also Table 3) |  |
| Months since stroke – mean (SD), range | 9.6 (8.4), 4–38 |
| Simplified Modified Rankin (SmRS) $\geq 3$<br>(classed as “dependent”) n (%) | 14 (74) |
| **Communication and cognition** |  |
| FAST expression – median (IQR) | 5 (3–5) |
| QAB comprehension – median (IQR) | 47 (40–48) |
| TOMS cognitive impairment, n (%) |  |
| 4 | 3 (16) |
| 4.5 | 10 (53) |
| 5 | 6 (32) |
| TOMS aphasia activity, n (%) |  |
| 2.5 | 1 (5) |
| 3.5 | 2 (11) |
| 4 | 1 (5) |
| 4.5 | 6 (32) |
| 5 | 9 (47) |

**Table 3.** Stroke Survivor outcome completeness, distribution and comparisons.

|  |  |  | **N** | **Mean (SD)** | **Change from Baseline (95% CI)\*** |
| --- | --- | --- | --- | --- | --- |
|  | ONS4 - Wellbeing | Baseline | 19 | 24.2 (5.5) |  |
|  |  | Post Grp | 15 | 24.1 (8.7) | -0.1 (-4.4 to 4.2) |
|  |  | 3 mnth | 15 | 23.5 (5.5) | -0.7 (-4.1 to 2.7) |
|  | HADS Depression | Baseline | 19 | 7.0 (2.6) |  |
|  |  | Post Grp | 15 | 7.0 (3.9) | -1 (-1.8 to 1.7) |
|  |  | 3 mnth | 15 | 6.5 (3.7) | -0.6 (-2.0 to 0.8) |
|  | HADS Anxiety | Baseline | 19 | 7.1 (4.2) |  |
|  |  | Post Grp | 15 | 7.4 (4.8) | 0.7 (-1.8 to 3.1) |
|  |  | 3 mnth | 15 | 6.9 (4.4) | 0.2 (-1.9 to 2.3) |
|  | EQ-5D-5L Health Utility | Baseline | 19 | 0.64 (0.22) |  |
|  |  | Post Grp | 15 | 0.70 (0.20) | 0.07 (-0.05 to 0.18) |
|  |  | 3 mnth | 15 | 0.73 (0.21) | 0.09 (-0.02 to 0.20) |
|  | EQ-5D-5L VAS | Baseline | 19 | 59.2 (18.7) |  |
|  |  | Post Grp | 15 | 63.6 (22.3) | 0.3 (-11.6 to 12.2) |
|  |  | 3 mnth | 15 | 66.3 (17.6) | 3.00 (-6.8 to 12.8) |
|  |  |  | **N** | **Mean (SD)** | **Change from Post-group (95% CI)\*\*** |
| **PROMS not collected at baseline\*\*** | AAQ-ABI – Psychological Flexibility | Post Grp | 15 | 18.7 (7.6) |  |
|  |  | 3 mnth | 15 | 16.9 (6.7) | -1.9 (-5.4 to 1.6) |
|  | VQ Progress values | Post Grp | 15 | 19.1 (5.7) |  |
|  |  | 3 mnth | 15 | 19.0 (7.7) | -0.1 (-3.4 to 3.3) |
|  | VQ Obstruct values | Post Grp | 15 | 11.3 (5.2) |  |
|  |  | 3 mnth | 15 | 9.4 (6.7) | -1.9 (-4.7 to 0.8) |

\*Change scores are for those with valid data for each outcome. A higher score indicates a positive outcome in the ONS4, EQ-5D-5L VAS, AAQ-ABI, VQ Progress Values. A lower score indicates a positive outcome in the HADS Depression, HADS Anxiety, EQ-5D-5L Health Utility, VQ Obstruct Values. \*\*For PROMS that were not collected at baseline (AAQ-ABI & VQ) change from Post-group to 3 month is in the final column.

### Objective 1: Workforce upskilling for fidelity and safety

The 13 facilitators included occupational therapists, physiotherapists, rehabilitation assistants, nurses, an assistant psychologist, a Stroke Association coordinator, and two NHS peer support workers with lived experience of stroke (only Site D community stroke team employed peer support workers). Most facilitators had no prior experience of delivering ACT as an intervention. In interviews, facilitators emphasised that effective delivery depended more on cross-cutting skills than professional background:

> *“I don’t think it’s necessarily qualification… it’s the ability to facilitate and encourage conversation.” [1404,Facilitator]*

Attendance at the optional weekly clinical supervision varied between facilitators; however, facilitators described the *availability* of psychological oversight – alongside the initial training from a neuropsychologist - as important for safe delivery:

> *“If I was delivering this without having that kind of person to reach out to, I’m not sure I’d feel as safe… it does need to have some psychology oversight.” [1201, Facilitator]*

Facilitator-completed fidelity checklists indicated that delivery was broadly consistent with the manualised protocol. Across 104 specified session components, only 9 (9%) were reported as not ‘fully’ delivered. These were most commonly related to insufficient time for detailed discussion. Session duration was generally close to the planned 120 minutes but varied between groups (mean session length 83–115 minutes across sites), often reflecting group size and pacing. No intervention-related serious adverse events were reported.

In qualitative feedback, facilitators described structured scripts and resources as helpful in supporting consistent delivery, particularly early on:

> *“I stuck to [the scripts] like glue at first… … as the weeks went on, I felt able to go off script” [1203, Facilitator]*
>
> *“The resources are amazing… I feel very lucky.” [1301, Facilitator]*

### Objective 2: Reach, inclusion & retention of marginalised stroke survivors

Of the stroke survivors, 14 (74%) identified as White or White British and 5 (26%) were from minoritised ethnic backgrounds (see Table 2). Communication and cognitive screening indicated 10 participants (53%) demonstrated some level of communication difficulties. Several factors influenced inclusion and participation; for example, reasons for non-referral or decline included severe communication difficulty or language barriers.

In interviews, some stroke survivors (SS) described peer connection as a key motivation for joining:

> *“what I wanted to get out of it is to particularly meet other people that are in the same position so that you don’t feel quite so alone.” [2304,SS]*

However, some described feeling less well matched to their group as their age and ethnicity was different from the rest of the group: *“I felt like the diversity statistic.” [2108,SS].* Communication difficulties also affected engagement: *“I can understand… but I can’t… communicate what I wanted to say” [2202,SS]*.

Facilitators noted structural limitations within routine NHS services may affect inclusion:

> *“We can’t use translators for our groups either, so we’re automatically missing people and that is so stigmatising and isolating.” [1303, Facilitator]*

Recruitment and retention of survivors from minoritised ethnic backgrounds and with communication difficulties was feasible. As described above, recruitment relied on existing NHS caseloads, which facilitators noted may not always reflect the ethnic diversity of the wider stroke population. Stroke survivors with communication difficulties also reported some barriers to engagement with the intervention.

### Objective 3: Feasibility of outcome data collection

Remote collection of outcome data was feasible across planned timepoints. Descriptive summaries of scores are presented in Table 3. Although assessment of change was not a primary objective, pre–post change scores are reported to explore the potential suitability of these measures as candidate primary outcomes for a future trial, consistent with our approach in WAterS-1 [22]

Fifteen participants (79%) provided PROM data post-group end and three-month follow-up. Most participants completed measures independently online, with support from researchers required for two participants. Missingness was low with one missing data point on three measures respectively across the whole dataset. Completion rates were high across all measures, indicating that remote administration was acceptable and practical.

### Objective 4: Acceptability & perceived value of the intervention

Post-group feedback surveys indicated high acceptability among both stroke survivors and facilitators. In stroke survivor feedback surveys (n = 15), 11(73%) agreed or strongly agreed that they enjoyed the groups, 14 (93%) reported that online delivery was suitable, 13 (87%) indicated they would continue using strategies learned, and 14 (93%) said they would recommend the groups to others. Despite enjoying and planning to continue using the strategies learned, survey ratings of perceived value / benefit were more mixed, with 7/15 (47%) agreeing that the groups had helped them, 7/15 selecting neutral responses and 1/15 disagreeing.

In interviews, many stroke survivors described using specific strategies in daily life:

> *“Yeah, I thought it was pretty good. I mean the grounding and the sort of meditative stuff was pretty good. I liked a lot of the stuff… I think it’s called mindful noticing”* (2409,SS)

Others described broader shifts in perspective: *“this has made me think that I can change things a bit” (2401,SS).* However, not all participants perceived benefit: *“I didn’t learn anything from it that I can say has helped me at all” (2406,SS)*

Facilitator responses (n=13) were positive overall. Most agreed that the session content was clearly defined (n=11,85%), appropriately pitched (n=10,77%), and suitable for online delivery (n=10,77%). All 13 facilitators reported a good understanding of the intervention, and 10 (77%) felt confident delivering it.

Facilitators similarly described observing value for participants, particularly the opportunity for shared experiences:

> *“The patients liked it being in a group… they liked kind of like bouncing off each other listening to each other’s experiences” (1101, Facilitator)*

Facilitators also noted that engagement and benefit could vary within individuals:

> *“He didn’t connect with some of the analogies… but he did like the breathing… So it’s not so, it’s like he did take something from the group.” (1302, Facilitator)*

Online delivery was generally acceptable, with most stroke survivors reporting it was suitable. However, all participants described reduced opportunities for connection, with one facilitator reflecting that it was *“not a particularly open environment… you’re talking to a screen” (1201, Facilitator)*, and some participants noting that face-to-face groups offered a different social atmosphere.

Overall, both stroke survivors and facilitators broadly described WAterS-2 as acceptable and useful, while recognising variability in individual experience.

### Objective 5: Contextual and implementation factors

Qualitative interviews highlighted contextual factors likely to influence future delivery of WAterS-2 within routine services. Although the intervention was generally valued and seen to complement existing rehabilitation, facilitators consistently described challenges integrating an eight-session, two-hour intervention into already-stretched community stroke services. Some felt shorter, more condensed or sections of the intervention could fit current practice:

“In its current format, no. But if the format was slightly different or tweaked, then yes… I think ACT could be used, 100%… I would just I think I would tweak it [the intervention] slightly from what it is at the moment” [*1203, Facilitator*]

Recruitment, coordination and delivery were reported as time-intensive and dependent on local workforce capacity. Despite the study offering a remote delivery model, NHS sites limited participation to individuals already on team caseloads and in geographical catchment areas dictated by NHS commissioning boundaries.

Online delivery was viewed as both enabling and challenging for all participants. Some valued the accessibility and reduced travel burden:

“I think that just makes it more accessible for everybody else, because we… couldn’t all get together anyway.” [2404,SS]

However, others felt that virtual sessions limited informal interaction and group cohesion. Facilitators described additional practical barriers including technology difficulties, language and communication needs. Views differed regarding optimal timing post-stroke, with some participants preferring earlier input and others suggesting delivery several months into recovery.

Facilitators reported that the ACT framework expanded their “toolkit” of approaches that could be incorporated into routine practice. Perceived enablers included structured manuals, supervision access and manageable group sizes, while constraints included session length, online format and language or aphasia-related accessibility demands.

Stroke survivor participants expressed mixed views about the ending of the programme. Some felt it provided sufficient tools to continue independently: *“I think this leaves you with enough to work with to progress.” [P2408,SS]*. However, others described the ending as abrupt and wanted more guidance: *“What’s the follow up and what are the next steps?… it’s actually quite a brief period of time.” [P2208 SS].* Several participants also valued the connections developed and expressed interest in ongoing opportunities for group support: *“It would be nice to get some people that have suffered the same things… together.” [P2304 SS]*. This highlights how WAterS-2 could link with longer-term psychological and peer support after stroke. Participants did not differentiate the value of peer connection between that from group members or peer support workers.

Figure 2 presents a provisional logic model linking the resources and activities required to deliver WAterS-2 with its proposed mechanisms of change and intended outcomes [41]. This may guide future refinement and evaluation of WAterS-2.

**Figure 2.**
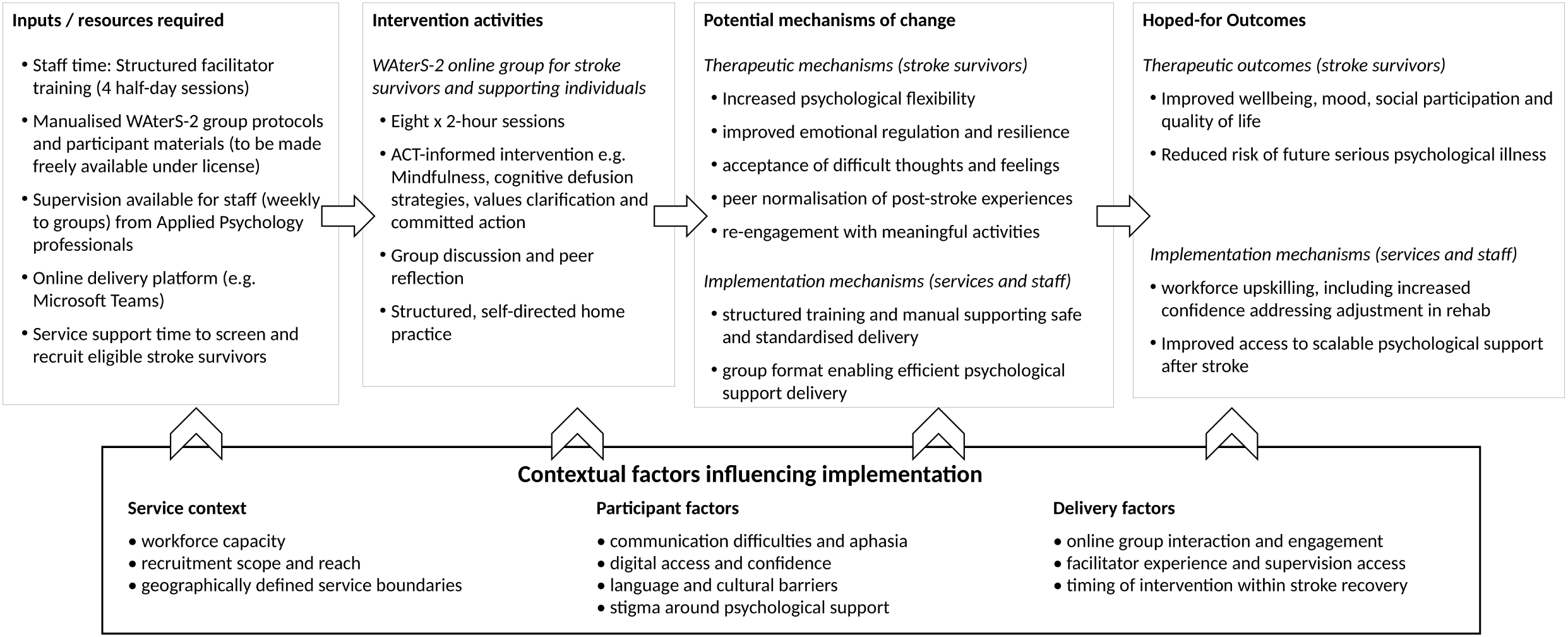
Provisional logic model for WAterS-2.

## Discussion

This multi-site feasibility study examined the delivery of an ACT-informed online group intervention within routine NHS stroke services, with a focus on workforce upskilling and inclusion of marginalised stroke survivors. Across four sites, facilitators from a range of professional backgrounds — including peer support workers — were able to deliver the programme with acceptable fidelity following structured training and clinical supervision. Stroke survivors with aphasia and from minoritised ethnic backgrounds were recruited and retained, though recruitment depended heavily on existing NHS caseloads. Remote outcome data collection was feasible, and both stroke survivors and facilitators broadly described the intervention as acceptable and useful. ACT is now recommended in national stroke guidelines [14] and RCT evidence for its efficacy is growing [12, 13], yet evidence for feasibility of NHS-based delivery through task-sharing models was lacking; these findings begin to address that translational gap, informing a provisional logic model and a foundation for designing a future pragmatic effectiveness-implementation trial.

We demonstrated that NHS staff who are not applied psychologists — including peer support workers with lived experience of stroke — could deliver an ACT-informed intervention with acceptable fidelity. WAterS-2 is a psychoeducational group intervention, sitting within a low-to-medium intensity tier of a matched care model; it is not intended as a replacement for specialist psychological therapy for individuals with complex or severe presentations [14, 42]. This is consistent with global evidence and WHO guidance on task-sharing models, showing structured training and supervision can enable non-specialist practitioners to deliver psychological interventions effectively [16, 18]. Psychological supervision appeared critical for safe delivery and should be treated as a core component of any implementation approach. Peer support (from group members or peer support workers) may add additional value also: evidence suggests that shared lived experience can enhance both engagement and credibility [43].

The study was constrained by wider NHS service structures. Facilitators consistently reported that community stroke teams were overstretched, so delivering an eight-session, two-hour intervention alongside existing care was challenging. However, facilitators described ACT as expanding their everyday “toolkit”, potentially giving them new ways to support stroke survivors’ psychological adjustment beyond the intervention itself. Crucially, delivery was bound by geographically-defined commissioning areas, despite the remote format. WAterS-1 utilised third sector delivery where national cross-boundary delivery was possible [22]. Future work may replicate this and/or explore peer-led or other cross-site delivery models to better fulfil the reach that telerehabilitation offers [19, 20].

Recruitment and retention of stroke survivors from minoritised ethnic backgrounds and with communication difficulties was feasible. This is a strength given that these participants are under-represented or even indirectly excluded in stroke research [44-46]. However, recruitment was entirely dependent on clinical teams identifying from existing caseloads, and supplementary community routes were not utilised, primarily due to NHS duty-of-care concerns. Language barriers, limited technology access, stigma associated with psychological support (which is potentially higher in minoritised ethnic populations [47]) all contributed to screening out or decline. Referral source and staff discretion can affect who is enrolled in research [48, 49], and introduces risk of systematic under-representation. Several facilitators reflected that their caseloads may not accurately represent the ethnicities of the broader stroke population — a concern supported by evidence of persistent ethnic inequalities in NHS stroke care and rehabilitation [9, 50]. Achieving genuine equity of access will require referral pathway reform and active community engagement strategies, not just inclusive intervention design.

ACT does not aim to reduce symptoms; rather, it enhances psychological flexibility and values-based living, which mediate downstream wellbeing [51-53]. Future trials should therefore include mechanism-focused measures (e.g. the AAQ-ABI and VQ) at baseline to explore change and incorporate longer follow-up periods to evaluate whether any early changes in psychological flexibility translate into subsequent improvements in emotional wellbeing. This study was not powered to detect effectiveness, and outcome data should be interpreted accordingly.

Study strengths include multiple NHS sites and integrated quantitative and qualitative data through a process evaluation, enabling a rich account of feasibility and implementation factors. Embedded People and Communities Involvement throughout the research process further strengthens the validity and relevance of the findings. Limitations include a small sample, and a single-arm design. Intervention delivery was restricted to one group per site; limiting exploration of variability across and within teams. Follow-up was limited to three months, which may be insufficient to capture effects of ACT. Finally, while fidelity was assessed using facilitator-completed checklists — a method used in prior WAterS research [39] — independent assessment was not conducted.

## Conclusion

WAterS-2 is feasible and acceptable for delivery within routine NHS stroke services by a mixed, non-psychology workforce. We demonstrated it is possible to recruit and retain stroke survivors, including people with aphasia and from ethnic minorities that are typically under-represented in research, and that remote outcome data collection is practical. However, NHS workforce pressures and geographically-constrained referral systems limit the potential reach of telerehabilitation; achieving broader equity of access will require changes to referral pathways, not just the intervention itself.

Findings provide a foundation for a provisional logic model and a future pragmatic trial. Such a trial should: rigorously evaluate ACT outcomes for this population; include mechanism-focused measures at baseline with longer follow-up to capture delayed effects; and explore alternative workforce and delivery models — including peer-led, cross-site delivery, and non-NHS routes — to maximise reach. All intervention resources and implementation guidance will be made freely available to support others working in the critical area of equitable psychological support after stroke.

## Supporting information

Appendices: 1. Tidier description of waters; 2. Fidelity checklist example; 3. Qualitative interview topic guides

## Acknowledgements

We are grateful to the stroke survivors and supporting individuals who took part in this study, and to the facilitators and participating NHS community stroke services who made delivery possible. We thank the members of our Research Advisory Panel, whose lived experience and insight shaped every stage of this research. These include: Ann Bamford, Rudolph Edwards, Billy Ellison, Sanya Karim, Jav Rehman, Wendy Simms, Stephen Taylor. We also thank the community organisations and connectors who extended the reach of our involvement work. We also thank our Clinical Leads in WAterS-2: Dr Victoria Teggart, Dr Natalie Hampson, Dr Lorraine King. A final thanks to members of our independent Steering Committee and Study Management Group for their oversight of study quality and conduct.

## Author Contributions

EP conceived the study, secured funding, and led the design and conduct of the research as co-Chief Investigator with AB. VL supported the qualitative analysis and drafting of the manuscript with EP. KWN contributed to data collection and study coordination. SC advised on methodology and feasibility outcome analysis. NC and ST contributed to study design and interpretation of findings. ABa chaired the Research Advisory Panel and led the involvement of people and communities, contributing to study design, materials, interpretation, and dissemination. AB contributed to study design, supervision, and interpretation of findings. All authors critically revised the manuscript for important intellectual content and approved the final version. EP is the guarantor.

## Data Availability

Data are available upon reasonable request. De-identified quantitative data and the study documentation are available from the corresponding author upon reasonable request, subject to ethical approvals. Qualitative interview transcripts are not publicly available, as they could compromise participant anonymity, but illustrative extracts beyond those reported may be requested. The WAterS-2 intervention manual and materials will be made freely available under license.

## Appendices

1. Tidier description of waters
2. Fidelity checklist example
3. Qualitative interview topic guides

