## Appendices: 1. Tidier description of waters; 2. Fidelity checklist example; 3. Qualitative interview topic guides for "Wellbeing After Stroke-2 (WAterS-2): a feasibility study with process evaluation exploring inclusive, accessible, online psychological support after stroke"

1. TIDIER description of waters
2. Fidelity checklist example
3. Qualitative interview topic guides

APPENDIX 1 – TIDER description of intervention [1] and session outline

| **Item** | **Description** |
|---|---|
| **Brief name** | **Wellbeing After Stroke–2 (WAterS-2)**: An 8-week, remotely delivered, Acceptance and Commitment Therapy (ACT)-informed group intervention for stroke survivors. |
| **Why (rationale, theory, or goal)** | WAterS-2 is grounded in the psychological flexibility model of ACT. It supports stroke survivors to adapt to life post-stroke by learning to be *OPEN* (making room for difficult internal experiences such as fatigue, anxiety, or frustration), *AWARE* (being present and mindful), and *ENGAGED* (taking meaningful, values-based action despite ongoing barriers). The overarching goal is to enhance wellbeing and psychological adjustment after stroke by supporting participants to live in alignment with their values, rather than focusing solely on recovery of deficits. |
| **Who for / When** | **Stroke survivors:** Adults ≥ 18 years of age in the UK, ≥ 4 months post-stroke (no upper limit), self-identified unmet psychological adjustment needs, sufficient English proficiency, and internet access.<br>**Supporting individuals (***if requested by stroke survivors to support engagement***):** Adults ≥ 18 years supporting a WAterS-2 group participant, sufficient English.<br>**Group size:** 4–10 participants per cohort. |
| **What** | **Materials:**<br><br>**Clinical protocol:** A structured facilitator manual (v8, 10/04/25) detailing scripts, timings, ACT-consistent enquiry guidance, slide prompts, and page references to the participant handbook.<br>**Participant handbook:** Posted before Session 1. Includes accessible worksheets (values clarification, "choice points", "passengers on the bus", "stepping-stones"), psychoeducation sheets (forgetting, fatigue), wellbeing signposting, and space for home practice. |

| Item | Description |
| --- | --- |
| | **Slide decks:** Screen-shared PowerPoint slides accompany each session for psychoeducation and exercises.<br>**Audio guidance:** Facilitators deliver short grounding/breathing practices from scripted wording.<br><br>**Procedures**<br><br>Each 2-hour weekly MS Teams session (including a 20-min break) follows a consistent structure: (1) settling and check-in; (2) review of previous home practice; (3) stroke-related psychoeducation (e.g. fatigue, forgetting); (4) experiential ACT exercises; (5) reflection and next-week planning. The 8 sessions progress from orientation and values clarification to mindfulness, acceptance of difficult experiences, and committed action. Session 8 focuses on consolidating a personal "Doing what it takes after stroke" plan. |
| **Who provided** | Delivered by trained *non-psychologist* healthcare professionals familiar with stroke but not ACT specialists. Facilitators complete a 4-week online training programme led by ACT-experienced clinical neuropsychologists and the WAterS-2 lead researcher. Training covers the ACT model (OPEN, AWARE, ENGAGED), enquiry skills, group process, and use of scripts. Weekly 1-hr online group supervision is offered during group delivery to maintain fidelity and support facilitators. |
| **How (mode of delivery)** | Small-group sessions conducted synchronously via Microsoft Teams. Facilitators lead sessions 'live', sharing slides, guiding exercises, and inviting discussion. Participants use their printed handbooks to follow along and complete worksheets. Engagement is verbal and via chat; participants may use cameras/microphones flexibly to manage fatigue or communication difficulties. |
| **Where** | Entirely remote. Participants join from home using their own devices and internet access. Facilitators deliver from their workplace or home. No assistive software was required; accessibility was supported through pacing, repetition, and workbook materials. |

| Item | Description |
|---|---|
| **How much** | Eight weekly sessions, each lasting two hours (including a 20-min break). See below for outline of 8 sessions. Total intended contact ≈ 16 hours over 8 weeks. Home practice between sessions is encouraged but self-directed and values-based (e.g. short mindful noticing, observing “choice points”, practising “stepping-stones”). Between-session contact with facilitators / team was not encouraged but permissible for attendance or welfare coordination. |
| **How challenging** | Sessions are cognitively and emotionally demanding rather than physically challenging, requiring sustained attention and engagement with difficult material (e.g. acceptance of stroke-related losses, values clarification). Challenge was calibrated to individual capacity through flexible participation, regular breaks, and self-directed home practice. Explored through qualitative interviews and feedback surveys. |
| **Regression / Progression** | Session dosage was fixed at eight weekly two-hour sessions with no planned progression or regression of session number or duration. The challenge level of session content (see above) increased progressively across the programme, moving from foundational psychoeducation and values work in early sessions to mindfulness, acceptance of difficult experiences, and committed action planning in later sessions. Within-session adaptation of pacing, language, and participation demands was at facilitator discretion, guided by clinical judgement and weekly supervision. |
| **Personalisation** | • **Individual values:** Each participant identifies personally meaningful values and related small actions.<br>• **Accessibility:** Language is simple; facilitators encourage flexible participation (chat, pauses, breaks).<br>• **supporting individual involvement:** Participants may nominate a *supporting individual* (e.g. family member or friend) to help them attend or use technology but not to co-participate in exercises. |
| **Protocol Deviations** | None major reported following finalisation of Protocol v8 (10/04/25). Scripts and timings are presented as flexible guides to be adapted responsively for group safety and accessibility. |

| Item | Description |
|---|---|
| **How well (planned and actual)** | Two complementary methods are available: (a) facilitator self-completed *Session Fidelity Checklists* confirming core content delivery; (b) independent *ACT-FM* ratings of recorded sessions assessing delivery process (ACT-consistent stance). WAterS-1 utilised both methods; WAterS-2 utilised only method (a) and suggested 91% protocol item adherence. Weekly expert supervision supports fidelity maintenance. |
| **Harms** | Planned: Potential adverse consequences considered included psychological distress arising from engagement with emotionally difficult material, and social discomfort within the group setting. Facilitators were trained to recognise and respond to signs of distress, with welfare concerns escalated through weekly clinical supervision. Participants were signposted to additional support resources in the handbook, and safeguarding procedures were in place in line with the study protocol [2]<br><br>Actual: No adverse events were recorded during WAterS-2 delivery. No participants reported or disclosed harm related to participation in the intervention. |

*Overview of WAterS-2 session content and linkage to ACT processes*

| Session | Stroke-related theme | ACT process emphasis | Key content / exercises | Home practice focus |
|---|---|---|---|---|
| 1. Coming together after stroke | Forgetting and memory problems | Orientation, compassion, early values | Group agreements; normalising distress; “making room for forgetting”; values reflection | Identify supports; prepare space for sessions |
| 2. How can I live better after stroke? | Fatigue and pacing | Values clarification, choice points | Explore what matters; “towards/away” moves; energy management | Notice choice points; take one small “towards” action |
| 3. Noticing more | Cognitive load / communication barriers | Present-moment awareness | Mindful grounding; “dropping anchor” | Daily brief mindful noticing |
| 4. Becoming more present | Worry and rumination | Cognitive defusion | “I’m having the thought that…” practice | Continue noticing thoughts and feelings |
| 5. Common feelings | Emotional responses to stroke | Acceptance / willingness | Identify and externalise difficult feelings as “passengers on the bus” | Greet “passengers” and take one “towards” step anyway |
| 6. Working with feelings | Pain, sadness, fear | Sustained willingness, self-compassion | “Passengers on the bus” continued. Reflect on urges to withdraw vs persist; practising kindness | Daily mindful noticing; reflect on meaningful moments |
| 7. Doing what it takes | Building realistic routines | Committed, values-based action | “Stepping-stones” planning; graded goal examples | Draft personal valued-living plan |
| 8. Living Better After Stroke | Endings and sustaining change | Integration of all previous lessons | Review and consolidate “Doing what it takes” plans; closure | Implement plan; use supportive self-talk |

### APPENDIX 2 - Fidelity checklist example

This is an example - there were 8 fidelity checklists in total (one for each session)

**Session 1 - Coming together after stroke**

***Please remember:***

- *Complete this ASAP following the session*
- *Fill this out honestly. Remember this is for our learning, not to penalise you*
- *Yes = component **fully** delivered as per protocol*
- *No = component **not** delivered or **partially** delivered*

•

| **Date:** | **Start time:** | **End time:** |
|---|---|---|
| **Number of participants:** | | |

| **Component** | | **Delivered as planned?** | |
|---|---|---|---|
| **Beginning the session (50 mins)** | | | |
| A | Introduction to facilitator and participants | No☐ | Yes☐ |
| B | Outline of session one | No☐ | Yes☐ |
| C | Introduction to the Participant Handbook [handbook] | No☐ | Yes☐ |
| D | Introduce ACT and the aims of the course | No☐ | Yes☐ |
| E | Outline the structure of the programme [handbook] | No☐ | Yes☐ |
| **Group rules (15 mins)** | | | |
| F | Core rules / agreements [handbook] | No☐ | Yes☐ |
| G | Adequate opportunity to discuss any additional rules | No☐ | Yes☐ |
| **Ending the session (20 mins)** | | | |
| H | Summary of the session | No☐ | Yes☐ |
| I | Adequate opportunity for reflections & comments | No☐ | Yes☐ |
| J | Home practice [handbook] | No☐ | Yes☐ |
| **If you've answered 'No' for any components, please give your reasoning (use the component's letter to identify which component/s you are referring to):**<br>*Please type as much as you like* | | | |
| **2 hours (approximately) to deliver all the session components was:** | Too short☐ | About right☐ | Too long☐ |
| **Any other comments?**<br>*Please type as much as you like* | | | |

### APPENDIX 3 –Qualitative interview topic guides

*Two separate topic guides were used: one for stroke survivor participants and one for facilitators. Both were conducted post-intervention and explored experiences of participation or delivery, acceptability, and implementation considerations.*

**Stroke Survivor Topic Guide**

1. Why did you choose to take part in the groups, and what were you hoping to get out of it? (Motivation to join)
2. Did this feel like the right time in your post-stroke journey to take part? (Timing and readiness)
3. What stands out most about your overall experience of the group — positive or negative? (Overall experience)
4. How easy was it for you to take part and follow the content? Were any parts confusing or difficult? (Accessibility and understanding)
5. How helpful did you find the course overall? Did it make a difference to your life in any way? Were there particular parts you especially liked or found less useful? (Impact and usefulness)
6. Are there any tools or strategies from the course that you plan to keep using? How confident do you feel about using them? (Continued use of strategies)
7. How did you find doing the course online? What worked well and what did not? (Online delivery)
8. How helpful was the handbook, and how much home practice were you able to do? (Handbook and home practice)
9. How suitable were the number and length of sessions, and how did you find the group size? (Practicalities)
10. How did you find being part of a group of stroke survivors rather than having one-to-one sessions? What were the benefits or challenges? (Group format)
11. Did you feel welcome and included in the group? What could we have done to make the course suitable for a wider range of people? (Inclusion and accessibility)
12. Did you feel prepared for the end of the groups? Do you plan to look for other support for your wellbeing afterwards? (Endings and future support)
13. Is there anything we have not talked about that you would like to share, or any suggestions for improving the groups? (Final thoughts)

**Facilitator Topic Guide**

1. How did you find the WAterS-2 training overall? What parts were most useful, and were there any you found less helpful or difficult? (Training experience)

2. Did you feel the training prepared you to deliver the intervention, including for participants with communication difficulties, cognitive needs, or from minoritised ethnic communities? What else might have helped? (Training preparedness)
3. What skills or background do you think facilitators need to deliver this intervention effectively? (Facilitator characteristics)
4. How did you identify and approach stroke survivors to take part? What worked well, and were there any challenges — particularly around including people from minoritised ethnic backgrounds or with aphasia? (Recruitment and inclusion)
5. What do you see as the main purpose of the therapy, and do you think it was beneficial for the people who attended? (Views on purpose and value)
6. How did you feel about delivering in a group setting? What were the benefits and challenges, including for participants with diverse communication, cognitive or cultural needs? (Group delivery)
7. How did you find delivering the intervention online? What worked well and what were the downsides or challenges? (Remote delivery)
8. Tell me about your experience of clinical supervision during WAterS-2 — did you use it, and if so did you find it helpful? (Clinical supervision)
9. How do you feel an intervention like WAterS-2 might fit within your current stroke pathway? What role could psychology or other professionals play in supporting delivery? (Implementation and fit)
10. Would you recommend this intervention to other facilitators or to stroke survivors? What might encourage or discourage others from taking it up? (Recommendation and future use)
11. Is there anything else you would like to share about your experience of delivering WAterS-2? (Final reflections)
